# Protocol paper: A.I. Based STroke Risk fActor Classification and Treatment (ABSTRACT) study

**DOI:** 10.1101/2024.12.11.24318721

**Authors:** William Heseltine-Carp, Aishwarya Kasabe, Megan Courtman, Michael Allen, Adam Streeter, Mark Thurston, Hongrui Wang, Lucy Mcgavin, Emmanuel Ifeachor, Stephen Mullin

## Abstract

**Background:** Stroke is a leading cause of death and disability in the UK. Much of stroke management revolves around addressing risk factors with medications and lifestyle modification. However, 30% of those who suffer stroke have no known risk factors. Hence, there is need to better identify individuals who are at high risk of stroke, and particularly those where the benefit of treatment outweighs the risk.

ABSTRACT is a three phase study that looks to address this issue by (1) using artificial intelligence (AI) to predict stroke risk from routine hospital data, (2) to validate this model on external datasets, and (3) validate the ability to improve outcome by guiding clinical decision making. In this paper we focus on phase I of the project.

**Aims:** Phase I of this study has 4 main objectives

1. *To create four separate machine learning (ML) models to predict stroke risk from routine hospital data. One for CT/MRI/PMH brain data, one for ECG/echo/PMH data and one for laboratory test/PMH data*.
2. *To perform explainability analysis on these models to identify important and novel risk factors for stroke*
3. *To calibrate these models and align them with real world probabilities*
4. *To combine these models to ensemble stroke prediction model*

**Methods:** In this retrospective observational cohort study we will analyze data from 9155 stroke patients and 109,875 controls in southwest England. Stroke cases will be sourced from the SSNAP database and historical brain imaging (CT/MRI), ECG, echocardiography, laboratory tests, ultrasound and medical history will be obtained from hospital and GP records. These data will then be linked to form a single de-identified dataset of cases and controls. ML techniques will then be trained on these data to predict stroke risk and identify novel risk factors for stroke.

**Discussion:** This protocol paper outlines phase one of ABSTRACTs approach in creating a novel stroke risk prediction model by integrating multimodal data types such as routine brain imaging, ECG, echocardiography, and laboratory results. In particular we outline a bespoke data handling protocol in order to comply with UK ethical governance when processing large volumes of confidential data. We also discuss our strategy in cleaning and preparing data prior to ML algorithms to predict stroke

## Background

Stroke is a leading cause of disability and death in the UK[1]. It also represents a substantial economic burden, as the first year of stroke aftercare costs on average £23,000 and £12,000 each year thereafter[2].

In recent years there has been a global push for a focus on preventative healthcare and risk factor modification[3]. Several major risk factors for stroke exist, such as dyslipidemia, hypertension, diabetes, previous stroke/transient-ischaemic-attack (TIA) and atrial fibrillation (AF). Modification of these risk factors through either lifestyle or pharmaceutical means significantly reduce the risk of future stroke[4]. However, despite this the incidence of stroke remains high[5]. This presents two major challenges.

Firstly, current healthcare is resource restricted and many of those prescribed treatments suffer dangerous adverse effects, such as bleeding with antiplatelet treatment[6]. To optimise cost effectiveness and safety it is important to understand in which individuals the benefits of these treatments outweigh the risks.

Secondly, 30% of those who suffer stroke have no known risk factors for stroke[7]. A better understanding of stroke risk factors is therefore required, paving the way for novel treatment strategies.

Explainable machine learning (ML) offers solutions to these issues. ML is capable of simultaneously handling numerous data types and identifying non-linear relationships between variables, enabling the development of accurate prediction models. Automated feature selection also makes ML well fitted to identify novel risk factors for stroke[8].

Due to its complexity, ML requires large datasets to accurately train models[8]. With the integration of electronic hospital records into routine clinical care, these datasets are readily available, however must be acquired through strict protocol adhering current to ethico-legal frameworks[9].

The A.I. Based STroke Risk fActor Classification and Treatment (ABSTRACT) study is a 3 phase project that aims to utilise artificial intelligence (AI) in stroke prediction and prevention. Phase I looks to use AI to predict future risk of stroke from routine hospital data, phase II aims to validate this AI model on an external dataset, and phase III looks to perform a clinical trial evaluating how this model can inform clinical decision making.

In this article we outline phase I of the ABSTRACT study where we will construct a machine learning model to predict stroke, from routine hospital data. Specifically, the data that we will be using to train this prediction model includes historical computerised-tomography(CT)/magnetic-resonance-imaging (MRI) brain, electrocardiography (ECG), echocardiography, carotid ultrasonography (US), laboratory test and past-medical-history (PMH) data from approximately 9155 stroke patients and 109,875 controls. In this paper we outline the protocol for this study, along with measures to ensure adherence to current ethico-legal guidelines.

### Aims

Phase I of this study has 4 main objectives

1. *To create four separate ML models to predict stroke risk from routine hospital data. One for CT/MRI/PMH brain data, one for ECG/echo/PMH data and one for laboratory test/PMH data*.
2. *To perform explainability analysis on these models to identify important and novel risk factors for stroke*
3. *To calibrate these models and align them with real world probabilities*
4. *To combine these models to ensemble stroke prediction model*

## Methods

### Study setting

The ABSTRACT study is a retrospective observational cohort study of 9155 stroke cases and 109,875 controls in South West England. This study looks to use routinely collected hospital data to train machine learning algorithms to predict future risk of stroke, and identify any novel risk factors for stroke.

### Study overview

Historical stroke cases admitted to Derriford hospital in Devon from Jan 2013 to Dec 2024 will be identified from the Sentinel Stroke National Audit Programme (SSNAP) database[10], a mandated national database of clinical data on those admitted to hospitals in England with an acute stroke. These cases will then be pseudonymised and linked with relevant historical CT/MRI/ECG/US/PMH/echocardiogram/laboratory test data from electronic hospital and General-Practice (GP) records. A pseudonymised control group of non-stroke scans and investigations will also be compiled, which is described below.

Once the linked and pseudonymised dataset of cases and controls have been generated, they will be de-identified and released to the researchers at the University of Plymouth for analysis. To begin with, separate stroke risk prediction models will be generated for each of CT/MRI/PMH, ECG/echo/PMH and laboratory test/PMH data. These models will then be subsequently combined into an ensemble model for stroke risk prediction, as illustrated in figure

1.Each of these stages are outlined in detail throughout this methods section.

### Eligibility Criteria

#### Inclusion

Cases will be identified from the SSNAP database and included on the basis of having an ICD-10 diagnosis of TIA/Stroke[11].

Controls were selected based on having no prior history of TIA/stroke in their GP or hospital records.

#### Exclusion

Participants will be excluded if they are under the age of 18 or have registered to opt-out of the study.

### Study co-design and public and patient engagement

Given the ethical considerations of using large datasets of confidential clinical data to predict stroke risk using ML algorithms, we opted to co-design elements of the project with members of the general public and those with lived experience of stroke/TIA.

To do this we began by surveying the general public and those with lived experience of stroke/TIA on their opinions in acquiring large clinical datasets via an opt-out model of consent and applying artificial intelligence algorithms to these datasets to predict stroke. The results of this work are detailed elsewhere[11], but in brief, the opinion of 83 participants (34% with a history of stroke) were evaluated for their opinions on the following questions; “Is it acceptable to train AI to predict future risk of stroke from routine hospital data?”; “Is it acceptable to acquire and handle large patient datasets using an “opt-out” model of consent?” and “Is it acceptable for members both within and outside of the routine clinical care team to have access to these datasets?” This was done using a combination of 1:1 interviews, focus groups and online survey. Overall, participants were found to have a favorable opinion of the study protocol. Based on this feedback, several adjustments were made to the protocol, including an “opt-in register” and advertising the project, along with how to opt-out, via non-electronic means, such as posters in public places.

Following this, an independent study oversight committee was then formed consisting of stroke survivors and members of the general public. This committee will oversee the handling of data, dissemination of results and plans for future research and clinical trials.

### Sample size calculation

Since the models also look to identify novel biomarkers for stroke prediction, production of precise sample size calculations is problematic. Given the lifetime prevalence of stroke is approximately 30%[12], we aimed for a case:control ratio of 1:3 in order to best reflect the anticipated case/control composition of a real life investigation library and to also account for missing data and potential opt outs.

In order to estimate the number of controls, 100 stroke cases were selected at random and audited for the availability of brain CT and MRI, ECG, echocardiography, PMH and laboratory tests investigations. The fraction of cases that had the investigation available was multiplied by 9155 to give an estimate of the total number of stroke cases who have this investigation available. This value was then multiplied by 3 to give an estimate for the required number of controls (table 2). Following this calculation we arrived at a figure of 9155 cases and 109,875 controls.

### Participant recruitment and data linkage

These large sample size estimations highlight the importance of complying with data protection regulations and ensuring adequate de-identification of subjects prior to analysis. We have therefore devised a bespoke pipeline to acquire, handle, store and link clinical records with relevant investigations in a secure and confidential manner. This pipeline is illustrated in figure 2 and is outlined in brief in the following steps.

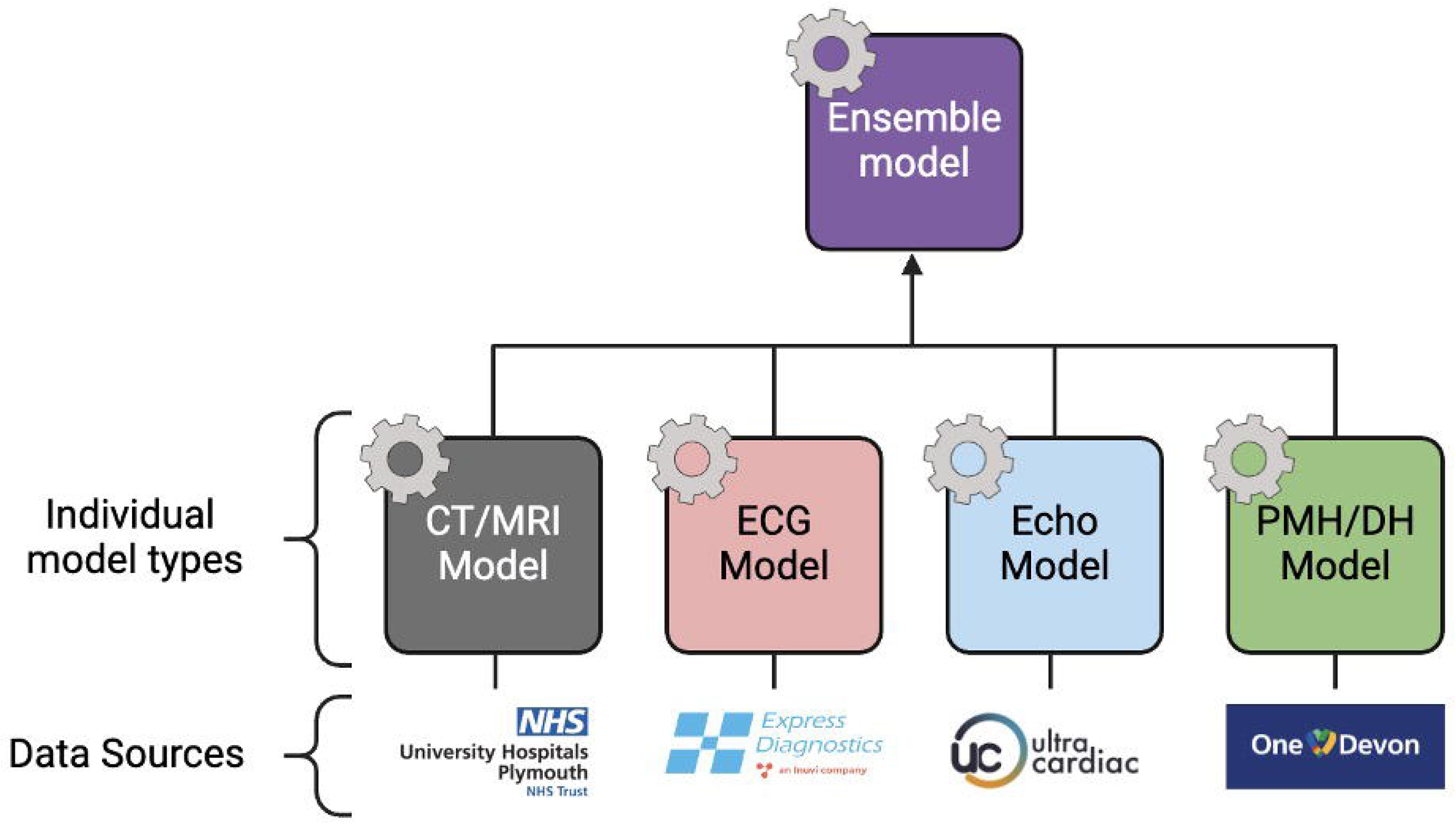

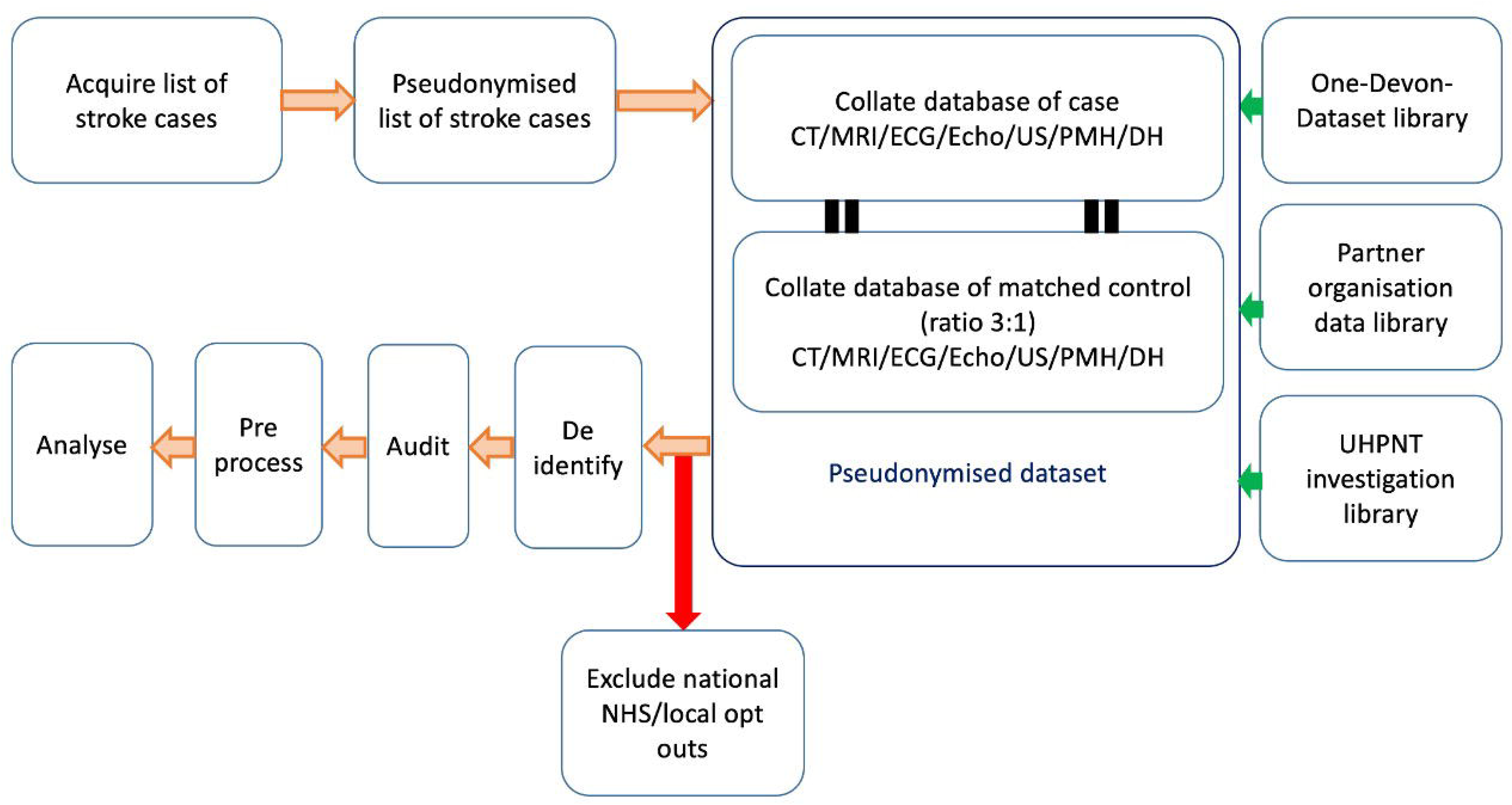

1)A list of subjects with stroke across University-Hospitals-Plymouth-NHS-Trusts (UHPNT) will be compiled from the SSNAP database by the UHPNT data manager, a member of the routine clinical care team who will act as an independent guardian of the data. They will hold an NHS contract and will be trained in the principles of Good Clinical Practice (GCP)[13] and General Data Protection Regulation (GDPR)[9].

2 This list will be pseudonymised by the UHPNT data manager and sent to the relevant routine clinical care teams (who will act as data source processors) for linkage with historical CT/MRI/ECG/echocardiogram/US/laboratory test data (using data such as NHS number, year of birth and the first four letters of the postcode). They will also generate a list of random controls with no history of TIA/stroke

Because each investigation will be held in a different database, specific data source processors will be required for each of CT/MRI, ECG, Echo/US, laboratory test and PMH data. The sources for each data type are outlined in table 1. Investigations were selected based on established or putative risk factors for stroke.

**Table 1:** Data definitions and sources.

| <u>Data type</u> | <u>Definition</u> | <u>Data source</u> |
| --- | --- | --- |
| Stroke cases | ICD-10 clinical diagnosis of stroke | SSNAP database, UHPNT electronic records (e.g. discharge summaries, patient letters) and one-devon-dataset. |
| CT/MRI brain | CT/MRI imaging of any region of the body, but with a focus on brain, head and neck imaging. CT/MRI data will include other modalities such as CT/MRI angiogram, venogram, plain film, contrast and gemstone spectral imaging. The data extracted will include data such as, pixel image data, request details (e.g. scanner used and hospital scan performed at) and the written report. | UHPNT electronic records, which are held on UHPNT databases, such as "patient archiving and communication system" (PACS), clinical record interactive search (CRIS) and iSOFT clinical manager (ICM). |
| ECG | Configurations such as 3 lead, 5 lead and 12 lead. This will also include data from portable devices, such as holter monitors and implantable loop recorders. | UHPNT electronic records, such as patient letters and ICM. Some of these data will also be obtained from express diagnostics (who are members of the routine clinical care team). |
| Echocardiography | This will include historical echocardiography and ultrasound data of any region of the body, with a particular focus on heart and carotid ultrasound imaging. This will include other relevant modalities, such as bubble study and doppler ultrasonography. | UHPNT electronic records, such as ICM, CRIS and PACS. Some of these data will also be obtained "ultracardiac" (who are members of the routine clinical care team). |
| Laboratory test data | Any biological sample sent to the haematology, biochemistry, cytology, pathology and/or microbiology department for analysis. In most cases this will relate to phlebotomy samples sent for tests such as full-blood-count, urea-and-electrolytes, C-reactive protein, liver function tests and lipids | UHPNT electronic records, such as ICM. |
| Past medical history | This will include historical diagnoses or encounters listed in the ICD-10, Patient demographics (such as date of birth, ethnicity, sex and postcode sector), and historical treatments for health conditions, including medical and surgical treatments. A detailed list of conditions included is listed in appendix 2. | SSNAP database, UHPNT electronic records and the one-devon-dataset. |
| Drug history | Historical prescriptions of any sort | SSNAP database, UHPNT electronic records and the one-devon-dataset. |
| **Table 1: Data definitions and sources.** |  |  |

**Table 2:** Estimates of total sample size based on an audit of 100 cases from the SSNAP database.

|  | Fraction who had investigation | Case estimate | Control estimate |
| --- | --- | --- | --- |
| CT | 1.0 | 1.0 x 9155 = 9155 | 9155 x 3 = 27,465 |
| MRI | 0.48 | 0.48 x 9155 = 4394 | 4394 x 3 = 13,182 |
| ambulatory ECG | 0.85 | 0.85 x 9155 = 7782 | 7782 x 3 = 23,364 |
| echocardiogram | 0.67 | 0.67 x 9155 = 6133 | 6133 x 3 = 18,399 |
| laboratory tests (at least FBC and U&E) | 1.0 | 1.0x9155 = 9155 | 9155 x 3 = 27,465 |
|  | Total sample size = Stroke cases + (CT controls + MRI controls + ambulatory ECG controls + echocardiogram controls + laboratory test controls + PMH/DH).<br>Total sample size = 9155 + (13,182 + 27,465 + 23,364 + 18,399 + 27,465)<br>Total stroke cases = 9155<br>Total Controls = 109,875<br>**Total sample size = 119,030** |  |  |

3) The pseudonymised, linked datasets of cases and controls from each of the data source processors will then be sent back to the UHPNT data manager for further linkage.

8).The UHPNT data manager will then combine each of the pseudonymised, linked datasets to a single pseudonymised, linked dataset of CT/MRI/ECG/Echo/US/laboratory test/PMH and DH data.

9) The UHPNT data manager will then compare cases and controls against a national and local opt-out database, removing them accordingly. They will then de-identify the final dataset before releasing it to researchers at the University of Plymouth for analysis. The details of the opt-out and de-identification processes are outlined below.

### Consent and Opt-out

It would be both impractical and introduce significant selection bias to seek individual consent. Hence, an opt-out model of consent will be used to acquire data. As mentioned, participants will have the option to opt-out at both a national and local level and will be removed from the database prior to de-idenitification and release of the dataset to University of Plymouth (UoP) researchers.

At the national level, participants will be cross referenced against the NHS England opt-out[14] database prior to data extraction, and removed accordingly.

At the local level, the study, and how to opt-out will be advertised via social media and posters in community areas, such as hospitals, GP practices and post offices. Subjects may opt out by emailing their details to an NHS data manager or logging their preference via the study website[11].

### De-identification

After removing opt-outs, the UHPNT data manager will de-identify the linked dataset before releasing it to UoP researchers for analysis.

To do this, NHS numbers will be removed from the linked database and replaced with computer generated randomised study numbers. A separate database will hold the linked NHS numbers and study number. This database will be held for the duration of the study and accessed only by the UHPNT data manager in order to allow ongoing linkage and resolution of data queries. It will be password protected and stored on an encrypted NHS server.

To ensure this process has adequately removed identifiers, automated testing will be run on the de-identified dataset to prove successful removal of identifiers. A manual audit of a representative sample of data will also be performed.

### Transfer of data

In any pipeline, stages involving transfer of data are vulnerable to leakage of sensitive information. We therefore designed our protocol to be particularly robust in the context of data transfer. The flow of data transfer is illustrated in figure 3. At each stage, data will be handled by the minimum number of qualified individuals.Transfer of identifiable stroke of case lists will be strictly carried out via encrypted email or file transfer and only within secure data environments of UHPNT or partner organisations delivering clinical care[15], [16]. CT/MRI/ECG/US/PMH/DH/echocardiogram/laboratory test data and relevant clinical data for analysis will be transferred to a university server either via direct upload or using an external hard drive, depending on capabilities and governance arrangements.

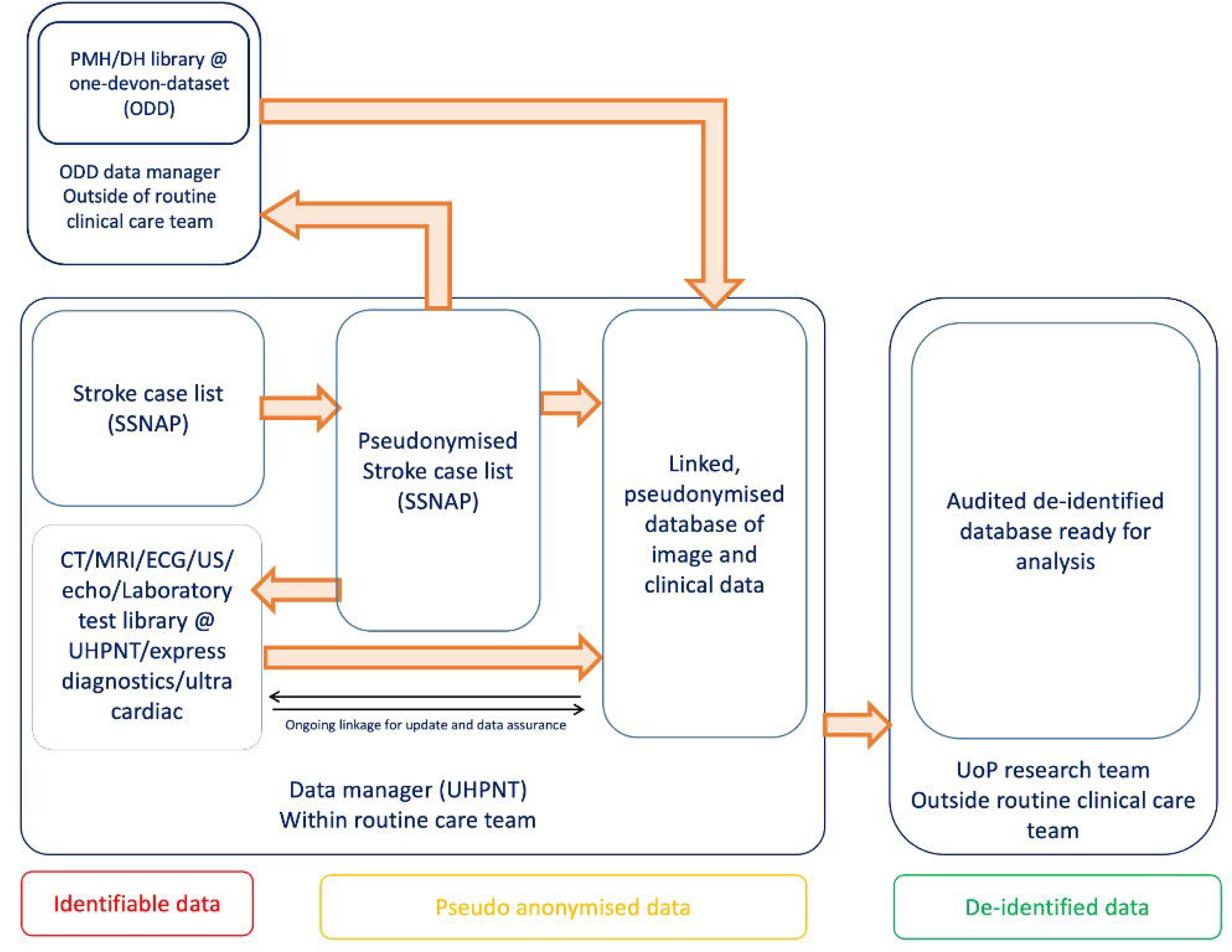

Most of this data will be sourced and handled by members within the routine clinical care team. This includes a portion of echocardiography and ECG data which will be obtained from (such as “express diagnostics” and “Ultra-Cardiac”), who perform these tests on behalf of the NHS. However, PMH and DH will be obtained from the One-Devon-Dataset (ODD) which are considered to fall outside of the routine clinical care team. In either instance the protocol has been designed to ensure that identifiable data is handled only by the UHPNT data manager (a member of the routine clinical care team), and only a pseudonymised list of cases a control is shared with data source processors for linkage purposes.

### Ethical approval

As discussed, CT/MRI/ECG/echo/US/laboratory test data will be acquired from UHPNT and third party organisations[15], [16] that work alongside UHPNT in carrying out routine clinical investigations. Data source processors at these organisations and UHPNT are considered part of the routine clinical care team, hence ethical approval was sought only from the research ethics committee (REC) for these data[17].

PMH and DH data will be acquired from ODD. The ODD is a collaboration between the Devon ICB and local authorities which holds data from organisations such as GP practices, NHS trusts, local authorities, and charities[18]. Data source processors at the ODD are therefore considered to be outside of the routine clinical care team, hence approval will be sought from the confidential advisory group (CAG)[19] for a section 251 waiver of common law obligations to confidentiality[20].

### End of study

At the end of the study any remaining patient identifiers will be destroyed and if feasible, an open-sourced, anonymised version of the dataset will be made publicly available as per UKRI guidance[21].

The end of study will be defined as follows:

1. Once the dataset has been fully analysed and the results of the study have been published in a peer reviewed journal.

We estimate this to be approximately February 2027.

### Analysis

Once de-identified, the linked dataset will be handed to UoP researchers for analysis. Separate ML stroke prediction models will be created for each of CT/MRI/PMH data, ECG/echo/PMH data, laboratory test/PMH data.

To begin with, the data for each model will be pre-processed, cleaned. In each case, data will be split into training and test datasets. Various models will then be trialled on each dataset before undergoing calibration and k-fold cross validation. Below we outline the intended model development strategy for each investigation type, prior to combining models to an ensemble stroke prediction model.

### Model development for each investigation type

#### For CT/MRI data

Data will be organised into common sequences. Images will be skull-stripped and registered to a standard MNI template[22]. Feature extraction techniques will be trialed to assess whether model performance is optimised by identification of features such as small vessel disease, infarcts and brain atrophy. Images will be cropped and resized to uniform dimensions prior to model input. Deep learning model architectures including convolutional neural networks and transformers will be trialed.

#### For ECG data

The Data signals will be subjected to several preprocessing techniques aimed at detecting noise and artefacts.Methods such as bandpass filtering, histogram analysis and template matching will be trialed for artefact detection. Additionally, the data will be examined for lead inversion to ensure proper peak detection.Following preprocessing, feature extraction methods will be applied to extract both beatwise and overall signal’s features. To optimize the feature set, feature selection techniques like Recursive Feature Elimination (RFE) and Sequential Feature Selection (SFS) techniques will be explored and evaluated. Various machine learning models like XGBoost, Recurrent Neural Networks (RNN) will be explored on the training dataset to evaluate the performance and determine the most suitable approach for this dataset.

#### For Echocardiogram data

Exploratory Data Analysis (EDA) will be performed to identify outliers, handle missing data, and remove uninformative features by analyzing correlations and evaluating the percentage of missing values. Imputation techniques such as mean, median, or K-nearest neighbors (KNN) will be applied for numerical data, while a separate category like ‘unknown’ will be created to handle missing values in categorical data. Performance metrics including Area Under the Receiver Operator Curve (AUROC) and accuracy will be used to ascertain model performance.

Several preprocessing steps will be applied for text-based features, including text cleaning (lowercase conversion, removal of stop words, punctuation, and special characters) and text vectorization (e.g., Term Frequency-Inverse Document Frequency (TF-IDF) and BERT embeddings) to transform text into numerical format. Feature selection techniques may also be explored before feeding the data to the model. Various machine learning models like XGBoost will be explored on the training dataset to evaluate the performance and determine the most suitable approach for this dataset.

#### For laboratory test data

To begin with data within 3 days or after the date of stroke will be excluded to avoid data leakage. Categorical and continuous variables will be manually defined and where appropriate encoded and converted to numeric data.

Data will then be standardized and explored graphically using histogram and distribution curves. A curve of best fit of stroke onset date and historical readings for each variable will also be plotted to visualise the temporal relationship between variables and stroke onset date. Packages such as Tsfresh[23] and Kats[25] will be used to engineer relevant time series features.

After relevant features have been identified, data will then be aggregated into weekly time intervals using the mode for categorical data and median for continuous data. Temporal data will also be standardized against age.

Techniques such as synthetic-minority-oversampling (SMOTE) and positive weight scaling will then be applied to the dataset to address imbalancing. Data will then be split into training and test datasets and datasets, which will then be used to train a variety of models such as XGboost, recurrent-neural-network, long-short-term-memory-models and support-vector-machine. Features selection will be refined using recursive-feature-elimination and tuning of hyperparameters.

#### Ensemble model development

Models trained for different investigations will then be combined into an ensemble model. The choice of ensembling technique will depend on the architectures and performances of the base models. Ensembling techniques that might be trialed include stacking, averaging, bagging (e.g. random forests) and boosting (e.g. AdaBoost or Gradient Boosting) [24]. Some of these techniques are better at combining diverse strengths of base models, whilst others are better at capturing complex relationships. As with the base models, the ensemble will be trained and assessed using k-fold cross-validation, and finally tested on a further holdout dataset (5% of total dataset). Explainability methods will be applied to explain ensemble predictions.

#### Missing Data

Inputs may be missing for some investigations for the ensemble model. The nature of the missing data will be analysed (to assess whether it is missing completely at random, missing at random, or missing not at random) in order to choose appropriate techniques for handling the missing data. Numerical data might be imputed using various methods. For unstructured data (such as imaging and ECG), ensemble weighting could be adjusted based on data availability. Sensitivity analysis will also be performed to identify significant bias caused by missing data.

#### Calibration

We shall check both individual models and the ensemble model for calibration to assess whether the distribution of the generated probability outputs reflect that of those who develop a future stroke. If necessary, we will improve the calibration by transforming model probability outputs (using methods such as Platt scaling, isotonic regression or beta calibration). Generated risk estimates will, if robust, be evaluated prospectively to predict future stroke in healthcare libraries.

#### Estimation of uncertainty

For both the individual models and the ensemble model, we will generate estimates of Epistemic and Aleatoric uncertainty around the predicted probability of stroke (as generated by our model) using the Dropout as a Bayesian Approximation method[26]. This method allows enhanced visibility of uncertainty generated by both a lack of available training data (Epistemic uncertainty) and that resulting from the lack of natural stochasticity of observations (Aleatoric uncertainty). This will allow both assessment of the potential for further model optimisation and of inherent limitations of approach. In lay terms this will allow assessment of the reliability of the predictions generated, which can guide decisions on whether it is suitable for deployment in a clinical context.

#### Validation

10% of the data will be withheld for validation purposes. 5% will be held to validate the ML models for each investigation, and the other 5% to validate the ensemble model. If the models trained perform as expected, we intend to carry our external validation at trusts outside of UHPNT (requiring further ethical approval).

#### Model interpretation

The strategies employed for each mode and the ensemble model will vary depending on the model architecture and the data type. For models that are inherently interpretable (such as regression models and decision trees), we will identify variables of interest using internal measures of feature importance. For models that are not inherently interpretable (such as neural networks), we will use explainability methods such as Gradient-weighted Class Activation Mapping (Grad-CAM)[25] and Deep SHapley Additive exPlanations (SHAP)[26] to identify and visualise features which have positively or negatively contributed to model outputs. Incorrectly classified examples will be reviewed, identifying systematic errors and avenues for optimisation. Correctly classified examples will be used to understand areas of interest to the model.

## Discussion

ABSTRACT is a 3 phase project that is the first large-scale study that combines multimodal data types such as, brain imaging, ECG, echocardiography, laboratory test results, and past medical history into a single machine learning model to predict stroke risk. The primary goal of phase 1 is to develop a model capable of accurately predicting stroke risk using routinely collected hospital data. We will begin by developing and calibrating separate models for each of CT/MRI/PMH, ECG/echo/PMH and laboratory test/PMH data, before combining these models into an ensemble model.

The secondary goal of phase 1 is to identify novel risk factors for stroke. A supervised machine learning approach was chosen for this purpose due to its ability to handle complex multimodal datasets and to perform automated feature selection[8]. Some models may be intrinsically explainable, but for those that are not, explainability techniques such as SHAP will be employed.

The development of such a model could have significant clinical implications, particularly in identifying high risk individuals who may benefit from aggressive risk factor management and targeted drug treatments. This is especially relevant in achieving cost effectiveness in a resource restricted setting. Furthermore, treatments like anticoagulants and antiplatelets carry a risk of bleeding[6], making precise risk prediction essential for patient safety.

Given 30% of strokes are cryptogenic [7], our study may also be useful in identifying novel risk factors and validating putative risk factors for stroke. We hope that this will subsequently inform new drug and treatment targets in stroke.

## Conclusion

In summary, this protocol outlines an innovative approach in using routine hospital data to predict future risk of stroke. In particular we outline a bespoke data handling protocol in order to comply with GDPR and ethical governance when processing large volumes of confidential data. The ability to identify high-risk individuals and novel stroke risk factors could significantly impact clinical decision-making and stroke prevention. Although there are limitations related to the retrospective nature and regional focus of the data, we will look to perform further external validation, which is outlined below.

## Supporting information

title page

## Data Availability

On completion of the study an anonymised version of the dataset will be made publicly available.

## Future Directions

Once phase 1 is complete, we aim to externally validate the model on datasets outside of the Devon/Cornwall region in phase 2. We will also plan to further explore and validate any novel risk factors for stroke that the model identifies at this stage.

In phase 3, we will then look to perform a clinical trial evaluating the role of aggressive modification of known or known risk factors and/or drug treatments, (such as statin and antiplatelet therapy) in those the model identifies as being at high risk of stroke.

