## Appendix 1 for "Protocol paper: A.I. Based STroke Risk fActor Classification and Treatment (ABSTRACT) study"

**RESEARCH REFERENCE NUMBERS**

| **IRAS Number:** | 306246 |
| --- | --- |
| **SPONSOR Number:** | 2557 |
| **FUNDER Number:** | EP/T518153/1 (ESPRC) |

### **KEY STUDY CONTACTS**

| Chief Investigator | Dr Stephen Mullin, University of Plymouth, 01752764487, |
| --- | --- |
| Sponsor | Mrs Sarah Jones, University of Plymouth, |
| Funder(s) | Engineering and Physical Sciences Research Council  Medical Research Council (MRC) |
| Co-Investigators | Prof. Emmanuel Ifeachor,  Dr. Adam Streeter,  Dr. Mark Thurston,  Dr. Lucy McGavin,  Dr. William Heseltine-Carp  Megan Courtman, [megan.courtman@postgrad](about:blank).plymouth.ac.uk  Aishwarya Kasabe |
| Committees | Dr Stephen Mullin, trial management group chair  [stephen.mullin@plymouth](about:blank).ac.uk, 01752764487, |

**FUNDING AND SUPPORT IN KIND**

| **FUNDER(S)**  (Names and contact details of ALL organisations providing funding and/or support in kind for this study) | **FINANCIAL AND NON FINANCIAL SUPPORT GIVEN** |
| --- | --- |
| Engineering and Physical Sciences Research Council | EPSRC DTP and School PhD scholarship |
| National Institute for Health Research | NIHR Academic Clinical Lectureship (SM)  NIHR Academic Clinical Fellowship (WHC)  NIHR Research Associate funding (MT) |
| Medical Research Council (MRC) | Research Grant |

**PROTOCOL CONTRIBUTORS**

**Dr. Stephen Mullin** is an Associate professor in neurology and consultant neurologist who will provide oversight and supervision to the project.

**Prof. Emmanuel Ifeachor is** a Professor of intelligent electronics systems. He will oversee design/building of the AI pipeline.

**Dr. Adam Streeter** is a medical statistician at the Peninsula Medical School and a biostatistician at the University of Munster. He will lead the data analysis/matching strategy.

**Dr. Mark Thurston** is a consultant radiologist at UHPNT. Together with Dr McGavin, he will oversee construction of the scan compilation pipeline.

**Dr. Lucy McGavin** Consultant neuroradiologist at UHPNT. Together with Dr Thurston she will oversee construction of the scan compilation pipeline.

**Dr. William Heseltine-Carp** is an NIHR Neurology academic clinical fellow. He will be involved in protocol design, governance, model development and project write-up.

**Dr. Hongrui Wang** Senior clinical data scientist. He will act as the data manager for the project.

**Dr. Megan Courtman** is a postdoctoral researcher who will be working on the project and will have primary responsibility for the data analysis and compilation, in addition to supervision for the machine learning process.

**Aishwarya Kasabe** is a PhD student whose primary responsibility will be analysis and development of machine learning models for the ECG data and Echocardiogram reports.

Amendments

The chief investigator will propose all substantial amendments to the study sponsor. If an agreement is made an amendment will be made via the IRAS amendment portal to the relevant ethics board.

Dissemination

Results of the study will be published in a peer reviewed journal. A lay version of the report will also be disseminated in a format accessible to the journal public, such as social media, magazine and newspaper. This will be aided by ongoing input from the study oversight committee.
