## Supplementary material for "Protocol paper: A.I. Based STroke Risk fActor Classification and Treatment (ABSTRACT) study": title page

Site of research: ITTC building, Plymouth science park, Plymouth, PL68BX

First author:

William Heseltine-Carp MBBCh, BSc(hons), MRCP, University of Plymouth, Room N6, ITTC building, Plymouth science park, Plymouth, PL68BX., +44 7540837485

Second author:

Aishwarya Kasabe, University of Plymouth, N6, ITTC building, Plymouth science park, Plymouth, PL68BX. 01752764487

Co-authors:

Megan Courtman PhD, University of Plymouth, N6, ITTC building, Plymouth science park, Plymouth, PL68BX. Plymouth, Plymouth, UK PL4 8AA. 01752764487

Michael Allen PhD, University of Exeter, Medical School, St Lukes Campus, Heavitree Road. SC 2.30. Exeter, UK EX4 4QJ. 01392 726080

Adam Streeter PhD, University of Plymouth. N15, ITTC1, Plymouth Science Park, Plymouth, PL6 8BX, +44 1752 764203

Mark Thurston, University of Plymouth. N15, ITTC1, Plymouth Science Park, Plymouth, PL6 8BX,

Hongrui Wang PhD, University of Plymouth. N15, ITTC1, Plymouth Science Park, Plymouth, PL6 8BX,

Lucy Mcgavin, University of Plymouth. N15, ITTC1, Plymouth Science Park, Plymouth, PL6 8BX,

Emmanuel Ifeachor PhD, University of Plymouth. School of Engineering, Computing and Mathematics, University of Plymouth, Plymouth, UK PL4 8AA +44 1752 586241

Stephen Mullin, MRCP PhD, University of Plymouth, Room N6, ITTC building, Plymouth science park, Plymouth, PL68BX., 01752764487
